# Reassessing the epidemiology of *bla*_CTX-M-15_: Emergence of *E. coli* ST1193 and potential replacement of ST131

**DOI:** 10.64898/2026.08.27.26361291

**Authors:** Alan Elena, Degrâce Batantou Mabandza, Uli Klümper, Sébastien Breurec, Christophe Dagot, Thomas Berendonk

## Abstract

The global dissemination of antimicrobial resistance is increasingly driven by bacterial clones combining antimicrobial resistance with enhanced virulence and environmental adaptability. *Escherichia coli* sequence type 131 (ST131) has historically been regarded as a major disseminator of the extended-spectrum β-lactamase (ESBL) *bla*_CTX-M-15_. However, the emergence of *E. coli* ST1193 carrying *bla*_CTX-M-15_ may represent an ongoing shift in the epidemiology of this resistance determinant. Here, we investigated the prevalence, genomic characteristics, virulence and antimicrobial resistance potential of ST1193 in comparison with ST131. A total of 1,136 *E. coli* isolates were recovered from touristic and non-touristic environments, hospital-associated samples, and aircraft toilets in Guadeloupe. Isolates were whole-genome sequenced and analysed for antimicrobial resistance and virulence determinants. Additionally, publicly available genomic data comprising 1,215 *bla*_CTX-M-15_-positive ST131 and ST1193 isolates were analysed to assess temporal and geographical trends. ST1193 was significantly associated with aircraft-associated samples and exhibited a higher antimicrobial resistance gene burden than ST131, while maintaining a comparable virulence factor content. Analysis of publicly available genomes revealed similar temporal emergence patterns for *bla*_CTX-M-15_-positive ST1193 and ST131, with ST1193 showing a more recent distribution and a higher number of deposited isolates in recent years, consistent with a potential ongoing clonal replacement. Comparative genomic analysis identified numerous virulence and adaptation-associated genes shared between both sequence types, while ST1193 additionally carried distinct determinants, including components of the transmissible locus of stress tolerance. Furthermore, quinolone resistance-associated mutations were strongly linked to *bla*_CTX-M-15_ carriage, particularly among ST1193 isolates. Together, these findings identify *E. coli* ST1193 as an emerging high-risk clone with substantial potential for *bla*_CTX-M-15_ dissemination. Its association with aircraft-associated samples further highlights the potential role of air travel in long-distance transmission and underscores the need to reconsider current surveillance strategies focused predominantly on ST131.

## 1. Introduction

Bacteria associated infections are a cause of major concern in both human and veterinary health, causing an estimated of 7,7 million deaths and disabilities yearly and costing the health systems around the world billions of dollars^1^.

Bacterial strains with increased virulence usually aggravate this scenario. Virulence factors are facultative genes that enable a series of bacterial functions that ultimately contribute to bacterial fitness and adaptiveness to a hostile environment^2^. Factors that increase biofilm formation are extremely useful for overall survival: The synthesised glycopeptide matrix allows bacteria within to growth unaffected by chemicals in the environment like disinfectants or antibiotics. Additionally, such structure promotes the adhesion to artificial structures increasing bacterial resilience to physical cleaning^3^.

Additional virulence factors involved in the formation of siderophores, adhesines, fimbriae and flagella favour bacterial persistence in the environment but also invasion and survival in the host^4–6^.

The carriage of antimicrobial resistance genes (ARGs) puts an additional strain on healthcare systems, rendering antibiotic therapies useless and therefore requiring longer and more expensive treatments for successful outcomes^7^. Among the most relevant, there are those that affect β-lactams or fluoroquinolones due to their safety and high usage rates^8,9^.

The combination of both high-virulence and important ARGs carriage in one clone is a major event that can lead to the worldwide dissemination of certain resistance markers and their endemic establishment in virtually all environments. One of the most prominent examples of such an association in the last century is the pandemic spread of *bla*_CTX-M-15_, an extended spectrum β-lactamse (ESBL), by the highly successful *Escherichia coli* sequence type 131^10^. The impact of this combination has been widely documented and is regarded as one of the most important contributors to beta-lactam resistance both in the community and in the nosocomial setting^10–13^. *E. coli* ST131 carries major virulence determinants that allow it to attach to surfaces, for biofilms, resist environmental toxins and survive under resource limiting conditions^14,15^. The incorporation of an ESBL gave it an additional competitive advantage and was mostly the reason of its sudden burst of cases.

Even when physiological aptitude and competitive fitness are a key driver in the dissemination and establishment of relevant pathogens, they should be analysed taking into account the behaviour of the host^16^. During the last century, human migration, mostly related to tourism, has been on the rise, facilitated by the accessibility of air travel^17^. The risk of pathogen dissemination by tourism has been made more tangible after the COVID pandemic and is a factor that should not be overlooked in a globalised society^18^.

Understanding the epidemiology of pandemic ARG carrying clones is a key factor to better informed healthcare systems that allow quick responses and prevent economic and sanitary costs.

In this work we explore the physiological characteristics of *E. coli* ST 1193 and its potential to lead a clonal replacement in the dissemination of *bla*_CTX-M-15_ based on its virulence profile, worldwide reports and association to air travel.

## 2. Methods

### 2.1 Sample isolation

Sampling was carried out on the French overseas territory of Guadeloupe. The defined low- tourism area was the city of Le Gosier, while the considered touristic zone was Pointe-a- pitre. Clinical samples correspond to the Jarry University Hospital.

Samples from the low and high tourism area involved household/hotel blackwater effluent, untreated and treated wastewater. One litre of water was collected in replicates in sterile glass bottles and filtered through a 0,22 µm pore size membrane, these membranes were then incubated in coliform count agar (CCA) for the isolation of *E. coli*, or in CCA+ceftriaxone (CRO - 4 µg/ mL) for the selection of ESBL producing *E. coli*. Colonies compatible with *E. coli* were reisolated in LBA or LBA+CRO (4 µg/ mL) and identified via MALDI-TOF. Hospital-related samples involved untreated effluent and treated wastewater but also patient samples. In the case of water samples, these were treated as mentioned. On the other hand, patient samples were recovered according to the appropriate methods depending on the site of infection. Aircraft toilette samples were recovered from the collection tanks and treated the same way as water samples.

For all the isolates, DNA was extracted using Qiagen’s PowerSoil Pro kit according to the manufacturer’s instructions^19^.

### 2.2 Bioinformatic analysis

#### 2.2.1 Sample sequencing

The isolated DNA was sequenced using an Illumina NovaSeq 6000 sequencer and a 2×100 bp sequencing approach. The average sequencing depth was 150x.

#### 2.2.2 Sequence retrieval

For the global analysis of *E. coli* belonging to ST131 and ST1193 we accessed the Enterobase^20^ (accession date: 15.12.2025) database and downloaded all available records. Samples with embargoed data or missing metadata was not included in this study.

#### 2.2.3 Sequence assembly

Adapters were removed from raw reads using BBduk v 38.96^21^. The same tool was applied to quality trim the reads applying a quality cut-off of 30. Quality trimmed reads were checked using FastQC v 0.12.1^22^ and then assembled using Unicycler v 0.5.1^23^ with default settings. Assembly quality was assessed using Quast v 5.3.0^24^.

#### 2.2.4 ARG and virulence detection

For the *in silico* detection of antimicrobial resistance genes and virulence factors, assembled genomes were queried against the ResFinder^25^ or the *E. coli* VF^26^ database using BLAST+ v 2.15.0^27^ and an identity and coverage threshold of 98%. For the specific case of the transmissible locus of stress tolerance (tLST), the LHR1 of *E. coli* AW 1.7 (CP073626.1), the LHR2 from *E. coli* FAM21805 (KY416992.1) and LHRa from *E. coli* S43 (CP010237.1) were used as reference.

#### 2.2.5 Sequence type assignment

Assembled genomes were classified into their corresponding sequence type using Achtman’s MLST scheme and the mlst tool v2.22.0^28^.

#### 2.2.6 Genetic annotation and analysis

For the overall genomic content analysis, we first predicted and annotated the ORFs present in each isolate using Prokka v1.14.6 only keeping contigs larger than 200 bp. A gene presence-absence matrix for each ST was generated with all the predicted ORFs using PPanGGOLiN v2.2.6^29^. In total, 29.203 unique genes were compared. The association of one gene to both or one of the STs was assessed by calculating an odds-ratio in R^30^. *p*-values were corrected for multiple testing using the Bonferroni method.

### 2.3 Statistical analysis

Statistical evaluation for associations between STs and sample origin was carried out by applying a Chi-test (*X*) test from in R. Significance in the difference of ARG and virulence factor carriage per cell between sequence types was done using a Kruskal-Wallis test followed by Dunn’s test.

## 3. Results

With the aim to identify the potential role of human migration through tourism on the spread of clinically relevant bacterial pathogens, we recovered human and environmental samples from 4 defined sampling groups: a) Touristic region, b) non-touristic region, c) nosocomial and d) aircraft toilette collections. All of these samples were recovered in the French overseas territory of Guadeloupe (Sample origin and location is detailed in table S1). A total of 1136 *E. coli* isolates were recovered (n_Touristic_ = 248, n_Non-touristic_ = 137, n_Hospital_ = 507 and n_Aircraft_ = 244), sequenced and analysed bioinformatically.

One hundred and eleven unique antimicrobial resistance genes (ARGs) were distributed across the isolates (median 3 ARGs/isolate) out of which the most common was the ESBL coding *bla*_CTX-M-15_, present in almost 30% of the isolates (n=338). Such an abundance led to investigate the behaviour and epidemiologic characteristics of such isolates in depth.

First, we assessed the sequence types (STs) occurrence and proportion in each type of sample. We observed 58 unique sequence types among which most were single occurrences, we therefore grouped the STs into “most abundant” (i.e.: those that made up at least 4% of the sample) or “others” (making up less than 4% of the sample). The most abundant ST included well-known clones such as: ST131, ST10, ST38 and ST39. To account for potential sample size effect, we calculated the proportions of each of the most abundant STs and analysed their distribution in each sampling group (Figure 1).

**Figure 1:**
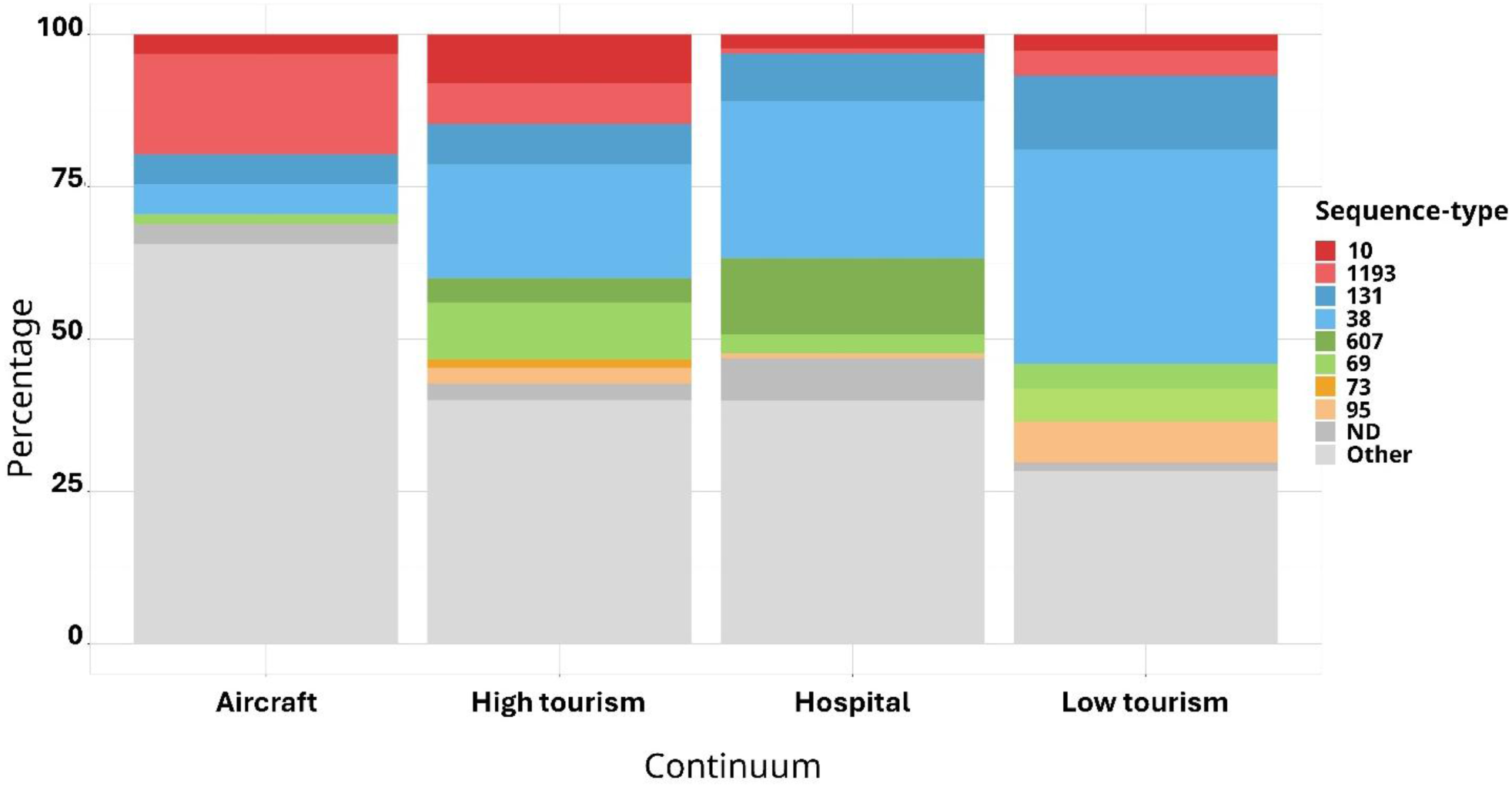
Proportion of major sequence types across samples by origin.

Based on recent epidemiology, the widespread presence and abundance of ST131 across all sample-types was not surprising. Instead, the sequence type 1193 stood out because of its ubiquity, but also due to its significant association with aircraft samples (*X-*test, residuals= 4,0349, *p*_Bonferroni_ = 0,01268). ST607 showed an important association with hospital samples (residuals= 4,2865, *p*_Bonferroni_ = 0,004211).

We proceeded to analyse the ARGs and virulence factor content in isolates belonging to the most frequent STs using the characteristics of ST131 as a benchmark for successful and high-risk behaviour. This decision was made based on the well-known characteristics of this clone that allowed it to act as a *bla*_CTX-M-15_ superspreader. Three of the analysed sequence types showed a higher ARG/cell content than ST131, being two of them significant (ST607 and ST1193). In contrast to this, when assessing the virulence factor content per cell, the representatives of ST607 showed a significantly lower carriage than ST131 while those isolates belonging to ST1193 showed very similar carriage (Figure 2).

**Figure 2:**
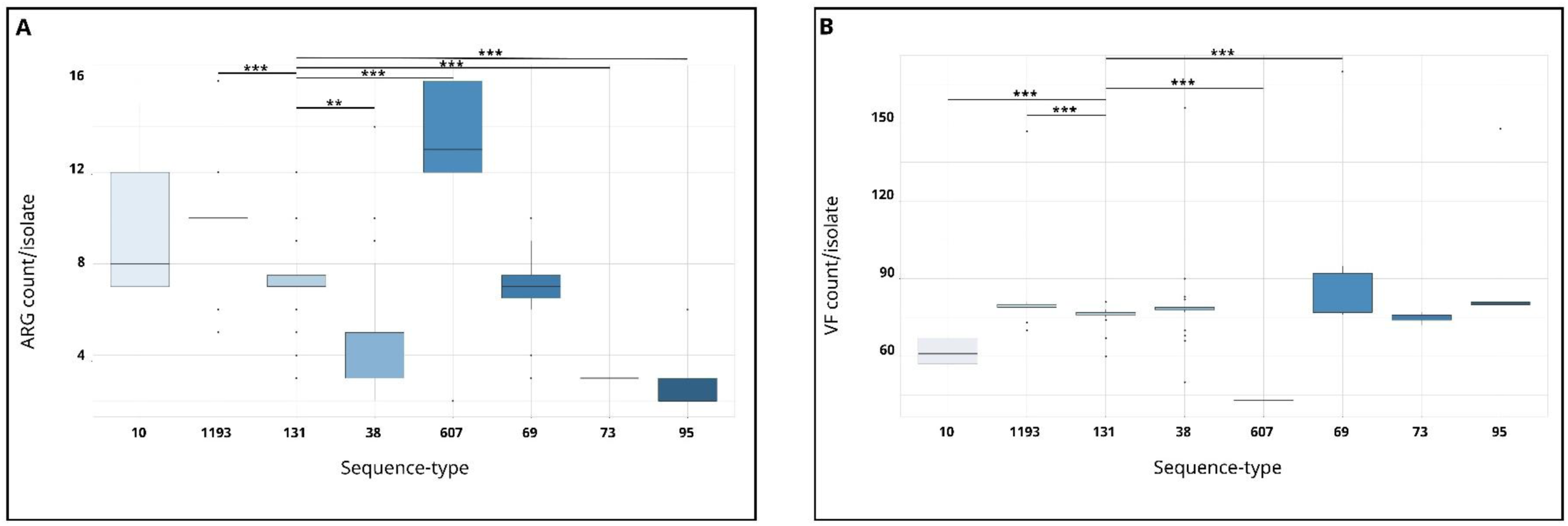
ARG (A) and virulence factor (B) carriage per isolate in each of the most common sequence types found in our samples. Significance is only shown with respect to ST131. *\* p<0.05, ** p=0.01, *** p<0.01*

These two characteristics, together with the aircraft association, suggested that ST1193 could be an equally relevant high-risk clone as ST131 with a potential for the dissemination of *bla*_CTX-M-15_ specially through means associated with human tourism.

### 3.1 Epidemiological shift: an ongoing process

Despite the mentioned concerns, *E. coli* ST131 is still regarded as the current most relevant *bla*_CTX-M-15_ spreader and driver of endemicity in scientific literature. Under this premise we wanted to investigate whether there was a potential ongoing clonal replacement favouring ST1193. For this, we accessed the Enterobase database and recovered all the publicly accessible genomic data from *E. coli* ST 131 (n=1962) and ST1193 (n=2044). We proceeded to screen for those isolates carrying *bla*_CTX-M-15_ using BLAST and the BLDB database. After discarding those isolates for which there was limited or no metadata we obtained a final working dataset of 1215 isolates (644 belonging to ST131 and 571 to ST1193). Samples had a diverse temporal and geographical background, eliminating potential outbreak biases.

In order to assess the potential epidemic behaviour, we explored the temporal dynamics of *bla*_CTX-M-15_ carriers of ST1193 and compared it to that of ST131. When analysing the occurrences per year, ST131 showed a bell-like shape showing a period of low occurrence in the early 2000s with a sudden and notorious increase reflecting the already documented ability of this clone to establish and successfully spread *bla*_CTX-M-15_. Notoriously, isolates belonging to ST1193 follow a similar shaped curve, suggesting a comparable phenomenon of high adaptation and successful spread. Such curve is shifted to the right compared to the one of ST131, meaning that their occurrence is closer in time than *bla*_CTX-M-15_ ST131. Even more, deposits of CTX-M-15 producing ST1193 isolates seem to be more abundant than those of ST131 in the latest years, suggesting that the significance of the association between ST1193 and CTX-M-15 is actively becoming higher than the ST131-CTX-M-15 association (Figure 3). It is important to note that the decrease observed in cases for both sequence types in 2023-2025 might not be an actual reflection of the epidemiological status, but rather a delay in sequencing and deposit from up-to-date data.

**Figure 3:**
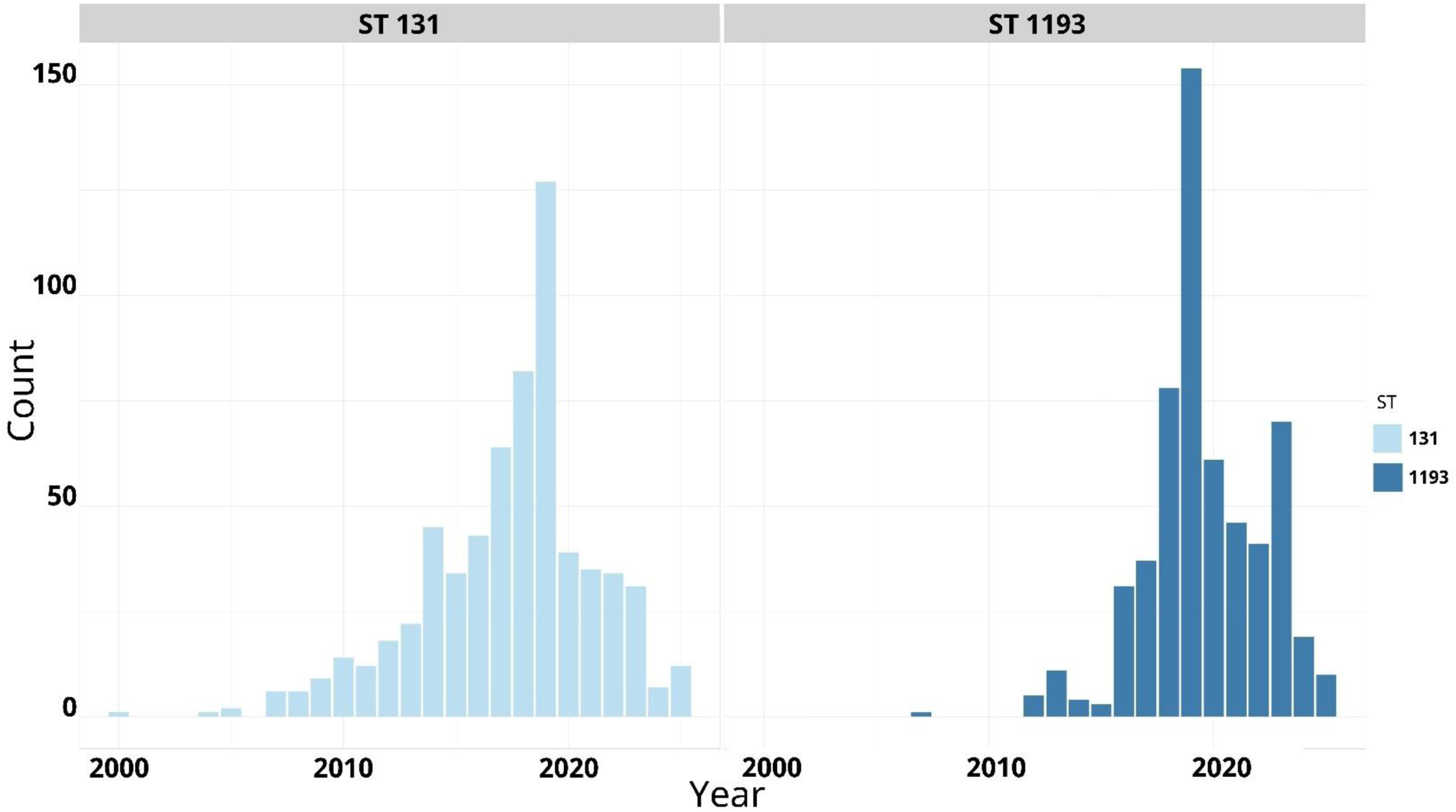
Occurrence of isolates harbouring *bla*_CTX-M-15_ belonging to ST131 (light blue) and ST1193 (dark blue) across the years. The data in this plot has been limited to the las 25 years for ease of visualisation.

### 3.2 Success potential

Having assessed the temporal dynamics of both clones, we sought to understand the underlying causes that allowed such a successful emergence of *E. coli* ST1193, and whether their genomic characteristics were enough for this clone to become as threatening as ST131. For this, we annotated all the genomes under study (including our own) using Prokka and generated a presence/absence matrix applying PPanGGOLiN. This process resulted in 29.203 identified genes. In order to test which genes were mostly associated to both or one or other ST we carried out an odds ratio assessment. For computational simplicity and to allow a more manageable and dedicated analysis we deeply analysed only those genes that were present in 95% of the cases in both STs (common genes) or only in 95% of the cases of one ST, but not in the other (Unique genes to one of both STs). Still, the full list of detected genes, odds ratio and *p*-value is available in Supplementary table 2 for future use.

Among the genes common to both STs we observed several virulence factors, with high potential to act as contributors to the success of ST131. These factors included the ferric-enterobactin uptake system *fepB/C/D/G*, *qseB* involved in quorum sensing, *adhE* a stabiliser of virulence, and *dmsA* a factor related to adherence and chlorine resistance (Table 1).

**Table 1:** Genes involved in virulence and drug resistance common *in E. coli* ST131 and ST1193.

| Sequence types 131 and 1193 |  |
| --- | --- |
| Gene | Relevance |
| *fepB/C/D/G* | Ferric-enterobactin uptake system<sup>31</sup> |
| *qseB* | Quorum sensing and flagellar activation<sup>32</sup> |
| *pilQ/hofQ* | Adhesin/Use of DNA as C source<sup>33</sup> |
| *sodB/C* | Oxidative stress defence<sup>34</sup> |
| *katG* | Oxidative stress response<sup>35</sup> |
| *msrP* | Oxidative stress protection<sup>36</sup> |
| *tus* complex | UPEC pathogenicity<sup>37</sup> |
| *dmsA* | Adherence under anaerobic conditions. Resistance to chlorine<sup>38</sup> |
| *glnA* | Survival in urinary tract<sup>39</sup> |
| *ahpC/F* | Oxidative stress response<sup>40</sup> |
| *adhE-2/P* | Stable expression of T3SS<sup>41</sup> |
| *bepA, bfr* | Increased virulence (Iron chelation) in starvation conditions<sup>42</sup> |
| *cfa* cluster | Motility and adhesion<sup>43</sup> |
| *fhuC/D* | Iron chelation in host<sup>44</sup> |
| *flgA/B1* | Flagellar formation<sup>45</sup> |
| *bla<sub>ampC</sub>* | Beta-lactamase<sup>46</sup> |
| *acrE/R/Z* | Multi-drug<sup>47</sup> |
| *mdfA* | Multi-drug<sup>48</sup> |
| *bcr-1* | Bicyclomycin resistance<sup>49</sup> |
| *sulA* | Sulfonamide resistance<sup>50</sup> |
| *tolC* | Multi-drug<sup>51</sup> |
| *abaF* | Fosfomycin resistance and biofilm production<sup>52</sup> |

Additionally, we observed that both STs carried almost ubiquitously genes that conferred resistance to antimicrobial compounds like disinfectants or antibiotics, among them: *sulA*, *bcr-1* and *abaF*.

As expected, *E. coli* belonging to ST131 had their own set of virulence factors. Genes allowing cell adhesion like *fim*, *pap* or *afa*, or biofilm formation like *marA* were uniquely present in this subset of organisms (Table 2).

**Table 2:** Virulence and chemical resistance genes associated only to *E. coli ST131*.

| Sequence type 131 |  |
| --- | --- |
| Gene | Relevance |
| *sadB* | Biofilm strengthenin<sup>53</sup> |
| *fimC* | Host cell adhesion<sup>54</sup> |
| *papC* | Host cell adhesion<sup>55</sup> |
| *afaD* | Invasin<sup>56</sup> |
| *fliC2* | Motility<sup>57</sup> |
| *yadC* | Adhesion and colonisation<sup>58</sup> |
| *mhpA/B/C/D/E/F/T* | Use of aromatic compounds. Potential survival in hostile environments<sup>59</sup> |
| *marA* | Stress, biofilm and multidrug efflux regulator<sup>60</sup> |
| *virB8* | Type-IV-SS Assembly<sup>61</sup> |
| *virB5* | Type-IV-SS Assembly<sup>62</sup> |
| *nfaA* | Host cell adhesion<sup>63</sup> |
| *mdtM* | Multidrug efflux<sup>64</sup> |
| *dfrD* | Confers resistance to trimethoprim<sup>65</sup> |

The absence of these genes in *E. coli* ST1193 doesn’t mean this clone has lower virulence potential, virulence factors only present in ST1193 isolates showed very similar functions, meaning that while not following the same identical strategies, both STs have the potential to be equally virulent (Table 3). As an additional virulence determinant, components of the transmissible locus for stress tolerance (tLST), specifically LHRa, LHR1 and LHR2, were detected only in representatives of ST1193. Even if these last determinants were only found in 4 isolates, their presence suggests the possibility of incorporation, stabilisation and dissemination of major adaptability characteristics.

**Table 3:** Virulence and chemical resistance markers associated to *E. coli* ST 1193.

| Sequence type 1193 |  |
| --- | --- |
| Gene | Relevance |
| *yadK/L/M/N/V* | Putative adhesion<sup>66</sup> |
| *virF* | Virulence regulon transcriptional activator (enteric pathogens)<sup>67</sup> |
| *bfpA* | Virulence regulator/Biofilm formation in enteric pathogens<sup>68,69</sup> |
| *espC* | Cytotoxicity mediated by TTSS<sup>70</sup> |
| *safA* | Quorum sensing/Adhesion<sup>71,72</sup> |
| *gspA1* | Type-II SS Component<sup>73</sup> |
| *pduA/B/C/D/E/F/L/U/V* | Propanediol utilization. Nutrient-limited conditions survival<sup>74</sup> |
| *ahpD* | Oxidative stress response<sup>75</sup> |
| *htrE* | Thermal tolerance/pillin production<sup>76</sup> |
| *mepA2* | Osmotic downshift resistance/Antibiotic decreased susceptibility<sup>77</sup> |
| *gatABC* | Niche competition/resistance to invasion by other bacteria<sup>78</sup> |
| *yfcO/P/Q/R/S* | Adhesion and colonisation<sup>79</sup> |
| *yraH/I/J/K* | Putative adhesion to bladder<sup>80</sup> |
| *tetC2, tetD3* | Resistance to tetracyclines<sup>81</sup> |
| *sat2* | Resistance to streptothricin<sup>82</sup> |
| *mccF* | Resistance to bacteriocins<sup>83</sup> |

## 4. Discussion

Based on retrieved sequences from public databases, the surge of ST1193 as a spreader of *bla*_CTX-M-15_ is not to be overlooked. The reports of ESBL producing isolates in the last 25 years follow a nearly identical pattern marking an identical dissemination potential. Two facts are worth highlighting: i) the number of deposited *bla*_CTX-M-15_ ST1193 is notoriously higher in the last years compared to *bla*_CTX-M-15_ ST131 suggesting a clonal replacement, and ii) the peak of *bla*_CTX-M-15_ ST1193 deposited sequences is shifted towards recent years, suggesting that the clonal replacement is still ongoing.

This dynamic is not reflected by the number of publications that involve ST1193, where ST131 is still studied and referred to as the main responsible for *bla*_CTX-M-15_ dissemination and invasion into a new niche^84–87^. Considering this, it is of crucial epidemiologic value to highlight a change in the epidemiology of this enzyme that might still be ongoing and that has gone mostly undetected.

Regarding the possibility of *E. coli* ST1193 to become a plausible clonal replacement for ST131 in *bla*_CTX-M-15_ epidemiology, our analyses indicate that both sequence types share virulence characteristics proper of high-risk clones, some of which have already been characterised as part of the success of ST131. Individually, isolates from ST1193 showed an extended set of virulence factors that can contribute even further to its adaptability in multiple settings, including the environment, hospitals and the human body. Even if low in frequency, the detection of tLST uniquely in isolates belonging to ST1193 signal a solid adaptability potential absent in isolates belonging to ST131.

Clonal exchanges of such magnitude are possible and have been documented previously with prominent cases like that of *Klebsiella pneumoniae* ST258 being replaced by hypervirulent *K. pneumoniae* after the surge of *bla*_KPC-3_^88,89^. Clonal replacements are multifactorial but often involve both bacteria and host lifestyle changes. In the present study, we found out that all the CTX-M-15 producing isolates belonging to ST1193 and ST131 carried some kind of mutation that conferred resistance to fluoroquinolones (*gyrA* S83A/S83L/D87N/D87Y, *parE* D475E/I529L or *parC* S57T/S80I/E84V), suggesting a strong association between both types of resistances. Even more interesting, we observed that non-CTX-M-15 producers belonging to ST131 didn’t have any kind of quinolone resistance related mutation in 64 cases, while this was true for only 5 isolates belonging to ST1193 (*Χ*=52,075 *p*<0,001). This shows that *E. coli* ST1193 might have a baseline advantage over *E. coli* ST131 and that the widespread consumption of β-lactams and fluoroquinolones (sometimes as combination therapy) might have been one of the driving forces involved in the proposed clonal replacement.

According solely to our data, isolates belonging to ST1193 were notoriously associated to samples recovered from aircraft toilettes, highlighting the ability of this clone to survive in such environments and therefore to spread over high distances in a short period of time. This sets ST1193 apart from ST131 and makes it a particularly dangerous clone in today’s society.

## 5. Conclusion

In this work, we provide specific data on the association between ST1193 and the dissemination of one of the most relevant ESBL worldwide, we also show time-resolved data that point out how ST1193 has surpassed ST131 as a *bla*_CTX-M-15_ spreader (on an occurrence basis) and is potentially continuing to do so currently. We additionally provide extensive data on genetic content of both clones with association values highly useful to explain the observed clonal dynamics. Additionally, we show based on our own data the higher potential of ST1193 to be disseminated via air travel.

A main concern regarding *bla*_CTX-M-15_ epidemiology is that despite the fact that *E. coli* ST1193 seems to be equally or more relevant than ST131, most works nowadays still take the latter as the most prominent clone^90–93^. As mentioned in the introduction, a proper understanding of the epidemiological status of highly relevant antimicrobial resistance genes and the clones responsible for their dissemination is a crucial factor for early detection and healthcare infrastructure in general. A manuscript by Pitout *et al.* is among the few to acknowledge the relevance of ST1193 as a high-risk clone^94^. In said work, it is mentioned that deeper understanding on the genetic capabilities that might contribute to this clone’s success are missing.

With this work, we expect to contribute to the visualisation of the risk posed by ST1193, allowing the medic and scientific community to take corresponding actions. In this sense, we are positive that the generated genetic data will bridge the gap mentioned by Pitout *et al.*, and be of further relevance for future studies involving the molecular characterisation of this emerging high-risk clone and its proper management.

## Data Availability

All data produced in the present study are available upon reasonable request to the authors

## 6. Acknowledgements

A.E. & T.U.B were supported through the PRESAGE project funded by the Bundesministerium für Forschung, Technologie und Raumfahrt (Grant number 02WAP1619) and by the RHUMARGE project funded by the Deutsche Forschungsgemenischaft (Grant number 544004729). U.K. & T.U.B were supported by the Explore-AMR project and the JPIAMR SEARCHER project funded by the German Bundesministerium für Forschung, Technologie und Raumfahrt under grant numbers 01DO2200 & 01KI24O4A. DBM and SB were supported by the Agence Nationale de la Recherche (ANR-20-AMRB-0001-01).

## Notes

### Competing Interest Statement

The authors have declared no competing interest.

